# *PRISMA2020*: an R package and Shiny app for producing PRISMA 2020-compliant flow diagrams, with interactivity for optimised digital transparency and Open Synthesis

**DOI:** 10.1101/2021.07.14.21260492

**Authors:** Neal R. Haddaway, Matthew J. Page, Chris C. Pritchard, Luke A. McGuinness

## Abstract

**Background:** Reporting standards, such as PRISMA aim to ensure that the methods and results of systematic reviews are described in sufficient detail to allow full transparency. Flow diagrams in evidence syntheses allow the reader to rapidly understand the core procedures used in a review and examine the attrition of irrelevant records throughout the review process. Recent research suggests that use of flow diagrams in systematic reviews is poor and of low quality and called for standardised templates to facilitate better reporting in flow diagrams. The increasing options for interactivity provided by the Internet gives us an opportunity to support easy-to-use evidence synthesis tools, and here we report on the development of tools for the production of PRISMA 2020-compliant systematic review flow diagrams.

**Methods and Findings:** We developed a free-to-use, Open Source R package and web-based Shiny app to allow users to design PRISMA flow diagrams for their own systematic reviews. Our tools allow users to produce standardised visualisations that transparently document the methods and results of a systematic review process in a variety of formats. In addition, we provide the opportunity to produce interactive, web-based flow diagrams (exported as HTML files), that allow readers to click on boxes of the diagram and navigate to further details on methods, results or data files. We provide an interactive example here; https://driscoll.ntu.ac.uk/prisma/.

**Conclusions:** We have developed a user-friendly suite of tools for producing PRISMA 2020-compliant flow diagrams for users with coding experience and, importantly, for users without prior experience in coding by making use of Shiny. These free-to-use tools will make it easier to produce clear and PRISMA 2020-compliant systematic review flow diagrams. Significantly, users can also produce interactive flow diagrams for the first time, allowing readers of their reviews to smoothly and swiftly explore and navigate to further details of the methods and results of a review. We believe these tools will increase use of PRISMA flow diagrams, improve the compliance and quality of flow diagrams, and facilitate strong science communication of the methods and results of systematic reviews by making use of interactivity. We encourage the systematic review community to make use of these tools, and provide feedback to streamline and improve their usability and efficiency.

## Introduction

### Evidence synthesis reporting standards

Evidence syntheses (e.g. systematic reviews and evidence maps) typically aim to reliably synthesise an evidence base, and are based on state-of-the-art methodologies designed to maximise comprehensiveness (or representativeness), procedural objectivity, and reproducibility, whilst minimising subjectivity and risk of bias (1, 2). Reproducibility is made possible through a high degree of transparency when reporting the planned or final methods used in a review protocol or final report. Reporting standards, such as PRISMA (Preferred Reporting Items for Systematic reviews and Meta-Analyses (3)) and ROSES (RepOrting standards for Systematic Evidence Syntheses (4)), aim to ensure that review methods and findings are described in sufficient detail.

In 2009, the PRISMA statement – a reporting guideline designed primarily for systematic reviews of health interventions – was released (3, 5). The guideline was developed by a consortium of systematic reviewers, methodologists and journal editors to address evidence of incomplete reporting in systematic reviews (6), with recommendations formed largely based on expert consensus obtained via Delphi surveys and consensus meetings. The PRISMA statement has been widely endorsed and adopted by journals, and evidence suggests use of the guideline is associated with more complete reporting of systematic reviews (7). However, to address the many innovations in methods for systematic reviews, changes in terminology, and new options to disseminate research evidence that have occurred since 2009, an update to the guideline (referred to now as PRISMA 2020 (8)) has recently occurred.

### Review flow diagrams

Flow diagrams in evidence syntheses allow the reader to rapidly understand the core procedures used in a review and examine the attrition of irrelevant records throughout the review process. The PRISMA flow diagram published in 2009 describes the sources, numbers and fates of all identified and screened records in a review (for more details, see the original flow diagram (3) and an update from 2014 (9)). A recent assessment of the quality and use of flow diagrams in systematic reviews found that only 50% of identified reviews made use of flow diagrams, with their quality generally being low and not significantly improving over time (quality defined by the presence of critical data on the flow of studies through a review (10)): as a result, the authors called for a standardised flow diagram template to improve reporting quality.

Several changes were made to the original PRISMA flow diagram in the 2020 update (8). The 2020 template: (i) recommends authors specify how many records were excluded before screening (e.g. because they were duplicate records that were removed, or marked as ineligible by automation tools); (ii) recommends authors specify how many full text reports were sought for retrieval and how many were not retrieved; (iii) gives authors the option to specify how many studies and reports included in a previous version of the review were carried over into the latest iteration of the review (if an updated review); and (iv) gives authors the option to illustrate the flow of records through the review as separated by type of source (e.g. bibliographic databases, websites, organisation and citation searching). Also, the phrase “studies included qualitative synthesis” has been replaced with “studies included in review”, given the former phrase has been incorrectly interpreted by some users as referring to syntheses of qualitative data. Furthermore, the recommendation to report in the flow diagram the number of studies included in quantitative synthesis (e.g. meta-analysis) has been removed, given a systematic review typically includes many quantitative syntheses, and the number of studies included in each varies (e.g. one meta-analysis might include 12 studies, another might include five).

### Transparency and Open Science in evidence syntheses

Broadly speaking, the Open Science movement aims to promote research integrity, experimental and analytical repeatability and full transparency, from project inception to publication and communication. Various definitions and frameworks for Open Science have been proposed (e.g. Open Data, Open Methods, Open Access, Open Source proposed by Kraker et al. (11), and 44 components by Knoth and Pontika (12)). In addition, the FAIR principles (Findability, Accessibility, Interoperability and Reusability (13)) aim to ensure that available data can be readily retrieved and used. In addition, licensing can be used to specify what can be done with the data once it has been accessed (14).

The application of Open Science principles to evidence synthesis has been explored by Haddaway (15), defined as *Open Synthesis*: the concept has since been expanded to cover 10 proposed components (<u>Open Synthesis Working Group 2020</u>). Open Synthesis is important and beneficial for a number of key reasons (15): 1) there is a need to be able to access and verify methods used in reviews and allow interrogation of the fate of each record in the review process; 2) in order to reduce research waste, data collected within a review should be made publicly accessible and readily reusable in replications, updates and overlapping reviews; 3) capacity building via learning-by-doing is facilitated by having access to machine readable data and code from a review.

### Interactivity and Web 2.0

Systematic review flow diagrams undoubtedly facilitate rapid comprehension of basic review methodology. However, they have far greater potential as a tool for communication and transparency when used not only as static graphics, but also as interactive ‘site maps’ for reviews. This is the essence of the concept ‘Web 2.0’; a rethinking of the internet as a tool for interactivity, rather than simply passive communication (16). Flow diagrams in their crudest sense consist of inputs, processes and outputs, with the ‘nodes’ (i.e. boxes) in a systematic review flow diagram containing summaries of the numbers of records included or excluded at each stage, and ‘edges’ (i.e. arrows) indicating the ‘flow’ or movement of records from information sources, through the screening stages of the review, to the final set of included studies. For each node, there is a rich set of information relating both to the methods used and the respective associated records: for example, the number of records excluded at full text eligibility screening are presented alongside a summary of the reasons for exclusion.

In a static review document, it may require substantial effort to determine the methods used to process records or the underlying records themselves. Indeed, the difficulty in locating the relevant information (particularly if stored in supplementary data) often hampers peer-review and editorial assessment. This is one of the key reasons that reporting standards require authors to specify the location of relevant information in review protocols or reports (e.g. see the PRISMA checklist; http://www.prisma-statement.org/PRISMAStatement/Checklist).

However, if we repurpose the flow diagram from a static element to an interactive ‘site map’ of the review, readers may immediately navigate to relevant information regarding review methods, inputs and outputs. Cross-linking between different elements of a review may help to facilitate the validation and assessment of systematic reviews and make it far easier to access and reuse their methods, data and code. Such interactivity could be achieved through hyperlinking within static digital files, such as PDF (portable document format) files, or through web-based visualisations that would facilitate updating or ‘living reviews’ (16).

Furthermore, by embedding and nesting relevant information behind an interactive visualisation such as a flow diagram, review authors could make use of a key concept in science communication: that of *simplification*. Simplification is a key principle in audio-visual science communication (17) and relies on prioritisation of information rather than ‘dumbing down’ (18). Extensive detail on the methods employed and on the reporting of information sources, data inputs and outputs could be accessed via hyperlinks, with core information placed front-and-centre. This layered or nested approach to science communication would allow the reader to choose how much and what type of information to view, rather than the linear format currently used across science publishing.

## Methods

### Objectives

This project had the following aims:

1. to develop a novel package for the R programming environment (19) for producing systematic review flow diagrams that conform to the latest update of the PRISMA statement (8);
2. to adapt this code and publish a free-to-use, web-based tool (a *Shiny app*) for producing publication-quality flow diagram figures without any necessary prior coding experience;
3. to allow users to produce interactive versions of the flow diagrams that include hyperlinks to specific web pages, files or document sections.

The project was produced collaboratively as part of the Evidence Synthesis Hackathon (https://www.eshackathon.org) using a combination of languages (R, DOT, HTML and JavaScript) with the aim of being provided to the public as a free and open source R package and Shiny app. The project code was published and managed on GitHub ((20); https://github.com/nealhaddaway/PRISMA2020) and the Shiny app is hosted on a subscription-based Shiny server paid for by the Stockholm Environment Institute (https://estech.shinyapps.io/prisma_flowdiagram). Code has been annotated and documented in line with coding best practices and to facilitate understanding and reuse. At the time of submission, the *PRISMA2020* package has been submitted to CRAN (the Comprehensive R Archive Network) for publication in their archive of R packages.

## Results

In the following pages, we summarise the functionality of the R package and Shiny app, providing a summary in lay terms, along with a more detailed description for the code-savvy (‘Code detail’ boxes). Functions are indicated by courier font, whilst packages are indicated by italics.

### The PRISMA2020 R package

#### Functionality

##### 1. Data import and cleaning

The data needed for the `PRISMA_flowdiagram()` function can be entered either directly as a set of numbers or R objects, but data upload can be facilitated by using a template comma separated value (CSV) file (see Table 1). We recommend the use of a CSV file as opposed to manually inputting numbers, as this allows for better reproducibility/transparency, as the underlying CSV can be shared. This file can be edited to a large extent and the edits incorporated into the text, numbers, hyperlinks and tooltips used to make the plot.

**Table 1.** Contents of the template CSV file for data upload.

| data | node | box | description | boxtext | tooltips | url | n |
| --- | --- | --- | --- | --- | --- | --- | --- |
| NA | node4 | prevstud | Grey title box; Previous studies | Previous studies | Grey title box; Previous studies | prevstud.html | xxx |
| previous_studies | node5 | box1 | Studies included in previous version of review | Studies included in previous version of review | Studies included in previous version of review | previous_studies.html | xxx |
| previous_reports | NA | box1 | Reports of studies included in previous version of review | Reports of studies included in previous version of review | NA | previous_reports.html | xxx |
| NA | node6 | newstud | Yellow title box; Identification of new studies via databases and registers | Identification of new studies via databases and registers | Yellow title box; Identification of new studies via databases and registers | newstud.html | xxx |
| database_results | node7 | box2 | Records identified from: Databases | Databases | Records identified from: Databases | database_results.html | xxx |
| register_results | NA | box2 | Records identified from: Registers | Registers | NA | NA | xxx |
| NA | node16 | othstud | Grey title box; Identification of new studies via other methods | Identification of new studies via other methods | Grey title box; Identification of new studies via other methods | othstud.html | xxx |
| website_results | node17 | box11 | Records identified from: Websites | Websites | Records identified from: Websites | website_results.html | xxx |
| organisation_results |  | box11 | Records identified from: Organisations | Organisations | NA | NA | xxx |
| citations_results | NA | box11 | Records identified from: Citation searching | Citation searching | NA | NA | xxx |
| duplicates | node8 | box3 | Duplicate records | Duplicate records | Duplicate records | duplicates.html | xxx |
| excluded_automatic | NA | box3 | Records marked as ineligible by automation tools | Records marked as ineligible by automation tools | NA | NA | xxx |
| excluded_other | NA | box3 | Records removed for other reasons | Records removed for other reasons | NA | NA | xxx |
| records_screened | node9 | box4 | Records screened (databases and registers) | Records screened | Records screened (databases and registers) | records_screened.html | xxx |
| records_excluded | node10 | box5 | Records excluded (databases and registers) | Records excluded | Records excluded (databases and registers) | records_excluded.html | xxx |
| dbr_sought_reports | node11 | box6 | Reports sought for retrieval (databases and registers) | Reports sought for retrieval | Reports sought for retrieval (databases and registers) | dbr_sought_reports.html | xxx |
| dbr_notretrieved_reports | node12 | box7 | Reports not retrieved (databases and registers) | Reports not retrieved | Reports not retrieved (databases and registers) | dbr_notretrieved_reports.html | xxx |
| other_sought_reports | node18 | box12 | Reports sought for retrieval (other) | Reports sought for retrieval | Reports sought for retrieval (other) | other_sought_reports.html | xxx |
| other_notretrieved_reports | node19 | box13 | Reports not retrieved (other) | Reports not retrieved | Reports not retrieved (other) | other_notretrieved_reports.html | xxx |
| dbr_assessed | node13 | box8 | Reports assessed for eligibility (databases and registers) | Reports assessed for eligibility | Reports assessed for eligibility (databases and registers) | dbr_assessed.html | xxx |
| dbr_excluded | node14 | box9 | Reports excluded (databases and registers): [separate reasons and numbers using ; e.g. Reason1, xxx; Reason2, xxx; Reason3, xxx | Reports excluded: | Reports excluded (databases and registers): [separate reasons and numbers using ; e.g. Reason1, xxx; Reason2, xxx; Reason3, xxx | dbrexcludedrecords.html | Reason1, xxx;<br>Reason2, xxx;<br>Reason3, xxx |
| other_assessed | node20 | box14 | Reports assessed for eligibility (other) | Reports assessed for eligibility | Reports assessed for eligibility (other) | other_assessed.html | xxx |
| other_excluded | node21 | box15 | Reports excluded (other): [separate reasons and numbers using ; e.g. Reason1, xxx; Reason2, xxx; Reason3, xxx | Reports excluded: | Reports excluded (other): [separate reasons and numbers using ; e.g. Reason1, xxx; Reason2, xxx; Reason3, xxx | other_excluded.html | Reason1, xxx;<br>Reason2, xxx;<br>Reason3, xxx |
| new_studies | node15 | box10 | New studies included in review | New studies included in review | New studies included in review | new_studies.html | xxx |
| new_reports | NA | box10 | Reports of new included studies | Reports of new included studies | NA | NA | xxx |
| total_studies | node22 | box16 | Total studies included in review | Total studies included in review | Total studies included in review | total_studies.html | xxx |
| total_reports | NA | box16 | Reports of total included studies | Reports of total included studies | NA | NA | xxx |
| identification | node1 | identification | Blue identification box | Identification | Blue identification box | identification.html | xxx |
| screening | node2 | screening | Blue screening box | Screening | Blue screening box | screening.html | xxx |
| included | node3 | included | Blue included box | Included | Blue included box | included.html | xxx |

The function `PRISMA_read()` reads in a template CSV file containing data to display in the flow diagram, including text contents, quantitative data (i.e. the number of records in each box), tooltips (i.e. the text that appears when the mouse hovers over a box), and hyperlinks for ‘on click’ functionality. The output is a list of named objects that can be read directly into `PRISMA_flowdiagram()`.

**Code detail:** The `PRISMA_read()` function uses text matching against a set of node (or box) names to assign the uploaded data to the appropriate box in the figure, for example:

~~~
   previous_studies <-data[grep(‘previous_studies’, data[,1]),]$n
~~~

##### 2. Creating a static flow diagram

The function `PRISMA_flowdiagram()` produces a PRISMA 2020-style flow diagram for systematic reviews. In summary, boxes are placed at specific locations across the graph, and they are automatically connected with arrows according to a specified set of connections.

**Code detail:** `PRISMA_flowdiagram()` uses the `grViz()` function from the *DiagrammeR* package (21) to plot a DOT graphic using layout = neato to explicitly place ‘nodes’ (boxes) at a particular location and `splines=‘ortho’` to specify axis-aligned edges are drawn between nodes. The label (including data) and tooltip for each node is read in within the main text of the function by using `paste()` to combine DOT strings and R objects.

Along with the text, data, tooltips and hyperlinks, users can specify whether to plot the ‘previous studies’ arm or the ‘other studies’ arm of the flow diagram by specifying these options within the `PRISMA_flowdiagram()` function.

In addition, the font, box fill, box line colour and line arrow head/tail can be altered as desired.

**Code detail:** Since text rotation is not supported in DOT or *DiagrammeR*, the vertical labels for the left-hand blue bars are added via JavaScript appending using the `appendContent()` and `onStaticRenderComplete()` functions from the htmlwidgets package (22) to append a block of JavaScript to the HTML output.

First, within the R code, a placeholder label is created consisting of a single whitespace. The JavaScript code uses the `document.getElementById()` to locate each of the blue bar nodes and replace the whitespace with the appropriate label. A CSS transform is applied to rotate the label by 90 degrees and the correct x and y coordinates for the new label are calculated based on their previous values. This means that the label location is adjusted based on the presence or absence of the ‘previous’ and ‘other’ arms and is able to withstand changes to the diagram format moving forward.

The function also includes the ability to plot an interactive version if the function parameter is set to ‘TRUE’, as described in Point 4, below.

The final plot output (see Figure 1) can be saved in a range of file formats (HTML, PDF, PNG, SVG, PS or WEBP).

**Figure 1.**
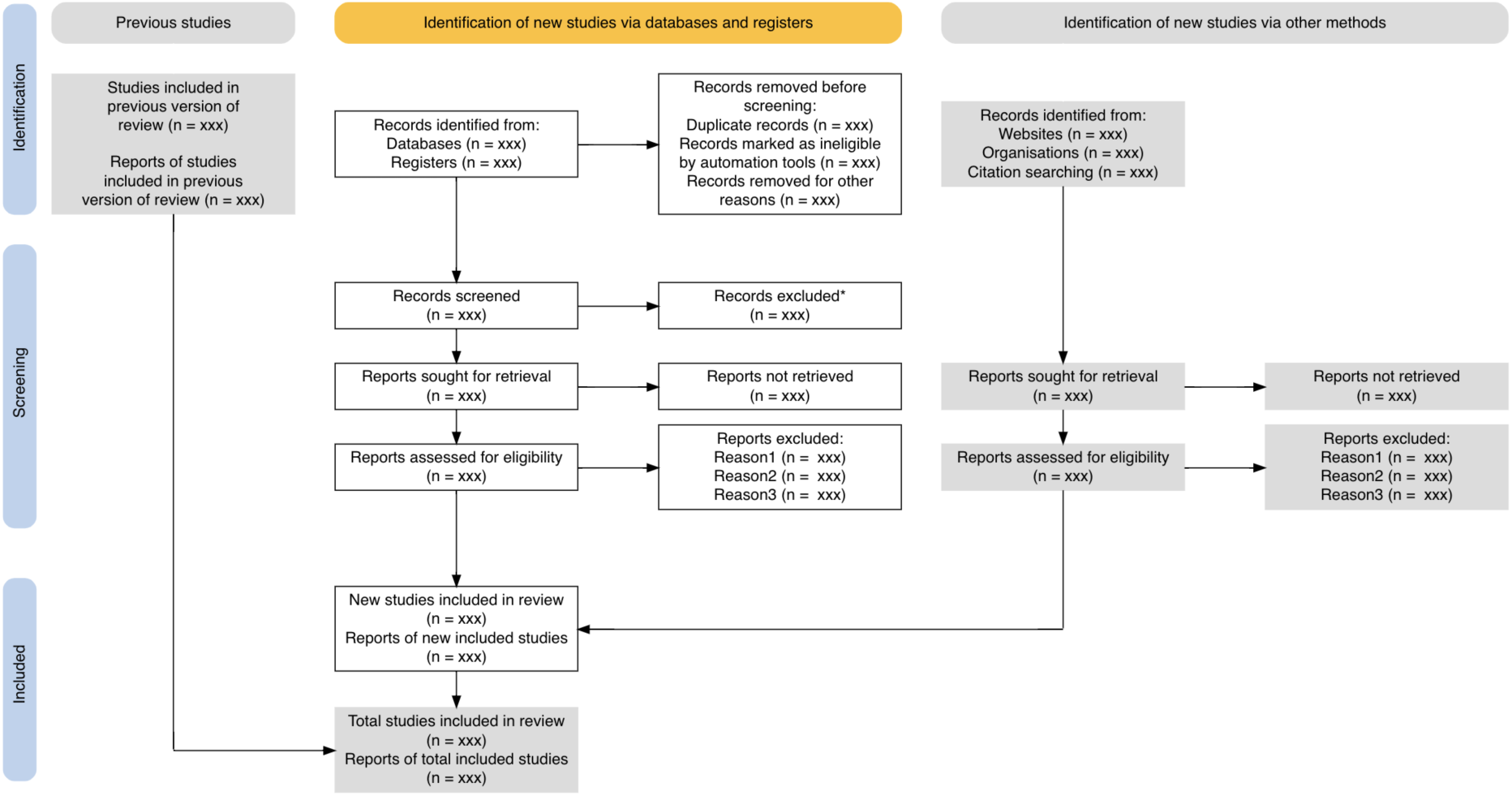
The full output plot from the `PRISMA_flowdiagram()` function.

##### 3. Creating an interactive flow diagram

Flow diagrams can be made interactive by specifying the additional parameter `interactive = TRUE` (this defaults to FALSE) in the PRISMA_flowdiagram() function. The resulting HTML output plot includes hyperlinks on click for each box, along with the tooltips specified in the main `PRISMA_flowdiagram()` function (see above).

**Code detail:** The internal function `PRISMA_interactive_()` uses the `prependContent()` and `onStaticRenderComplete()` functions from the *htmlwidgets* package (22) to prepend a block of JavaScript to the HTML output. This JavaScript identifies each node in turn using `getElementById(id)` and inserts an HTML anchor element carrying the relevant hyperlink for each node using the internal function `PRISMA_add_hyperlink_().`

##### 4. Saving the output as a file

The `PRISMA_save()` function allows for the flow diagram to be saved as a standalone HTML file (with interactivity preserved), or as a PDF, PNG, SVG, PS or WEBP file (without interactivity). This function takes the plot produced by `PRISMA_flowdiagram()` and saves the file. A default option for filename is provided, but this can be overridden along with the filetype which is calculated from the file extension by default.

**Code detail:** When saving as HTML, the `PRISMA_save()` function uses the `savewidget()` function from the *htmlwidgets* package (22). When saving as other formats, the internal function `PRISMA_gen_tmp_svg()` is used, this first uses `savewidget()` to create an HTML file in a temporary directory and then uses the various XML manipulation functions from the xml2 package (23) to step through the HTML, using xpath (24) to find the SVG embedded within the HTML.

As JavaScript is not supported in SVG files, the *xml2* package is again used to add a rotate transformation and programmatically alter the x and y coordinates to create the blue vertical labels. Following this, the temporary SVG is either copied to its final destination, or the rsvg (25) package is used to convert it into the desired output format.

### The Shiny app

Shiny is a package within the R environment that allows users to construct standalone web-based applications based on R functions (26). The ‘app’ can be interacted with by entering data, running functions with user-specified settings to plot figures, and downloading the resultant figures in a variety of formats.

The *PRISMA2020* Shiny app is available free-of-charge and can be found through the PRISMA website (http://prisma-statement.org/). The app landing page (the ‘Home’ tab) describes the app and its background, linking to the PRISMA website and PRISMA 2020 statement (8) (see Figure 2). Users can enter their data either by uploading an edited template CSV file, or by manually entering data in the ‘Create flow diagram’ tab. Once uploaded, users proceed to this tab to see the resultant figure.

**Figure 2.**
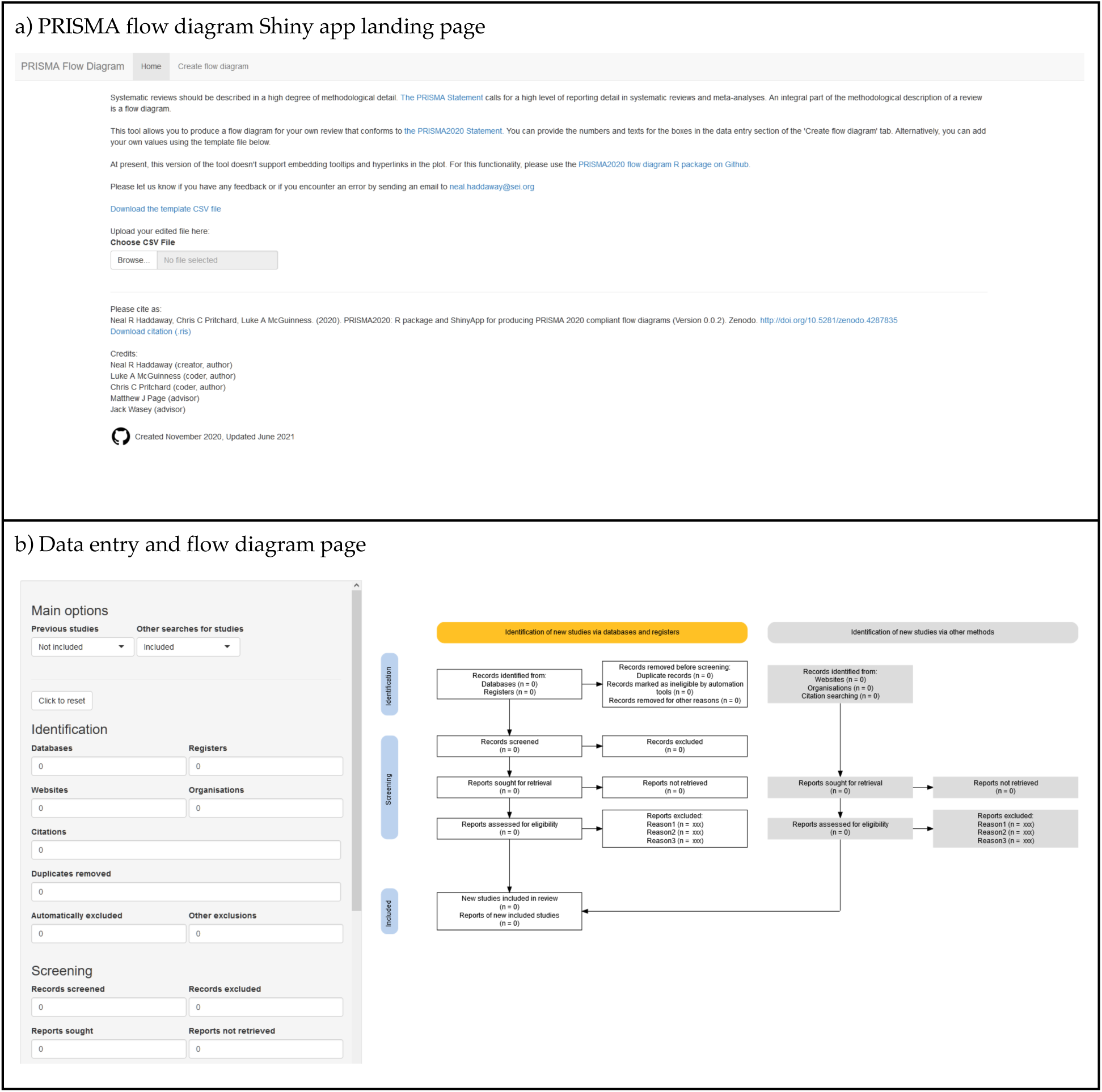
Screenshot of the *PRISMA2020* Shiny app a) landing page and b) data entry and diagram visualisation page.

On the ‘Create flow diagram’ tab users can specify whether to include the ‘previous studies’ and ‘other studies’ arms of the flow diagram using the check boxes. The resulting flow diagram (see Figure 3). The ‘Previous studies’ and ‘Other studies’ arms can be toggled on and off via the ‘Data upload’ tab and the plot responds reactively.

**Figure 3.**
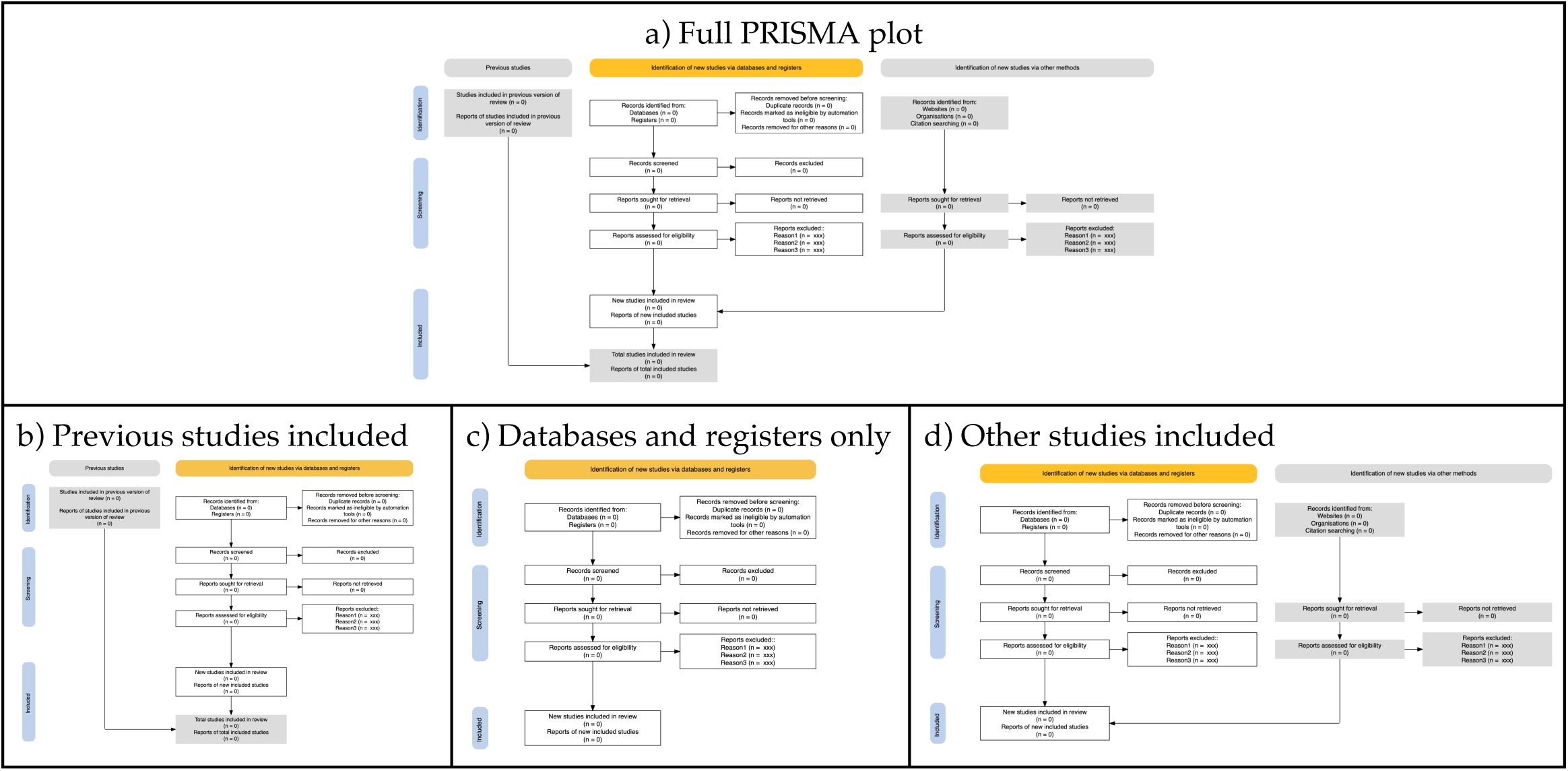
The possible layouts that can be obtained via the ‘previous studies’ and ‘other studies’ arms checkboxes. a) the full plot; b) other studies omitted; c) previous studies and other studies omitted; d) previous studies omitted.

Whilst interactivity isn’t possible using the Shiny app itself, the app allows for the download of an interactive HTML plot. The links themselves can only be customised through the upload of a custom CSV file, rather than through the web interface.

**Code detail:** Shiny does not support HTML appending or prepending (adding code before or after a given element) via *DiagrammeR*. Instead, a different method is used to label the blue nodes. The javascript file `labels.js` is included via a script tag inserted in the `<head>` area of the shiny HTML pages. This contains several functions, the `renderLabel()` function adds a label to the given node, in the same way as the JS appends in *DiagrammeR*.

The `createLabels()`function is called just before the plot is re-rendered, this registers a `MutationObserver` that waits for the nodes to be created. Once the nodes are visible to the DOM (Document Object Model, a programming interface for HTML), the `renderLabel()` function is called, once for each node, to add the labels.

### Interactivity

The interactivity here represents an additional step to cross link and host the interactive PRISMA flow diagram with the relevant texts and data. This obviously corresponds to additional effort on the behalf of review authors, but has clear benefits for transparency and communication.

Interactivity in the *PRISMA2020* flow diagrams is provided in 2 ways. Firstly, mouse-over tooltips appear as the user’s mouse is moved over a particular box. These popup boxes can contain user-specified text providing more information as desired. For example, a short elaboration of the numbers of text in each box in order to clarify meanings. Alternatively, tooltips can provide an explanation of the information that will be hyperlinked to on clicking. Secondly, the boxes can be given hyperlinks so that the user can follow a predetermined link. These links can be anchors within a document or webpage, or datafiles or web pages stored on external or local repositories (e.g. supplementary files on a data repository such as figshare or Zenodo).

As described above, this interactivity conforms to the principle of science communication simplicity by prioritising information provision hierarchically (i.e. showing critical information first, with further details accessible on interrogation). Tooltips provide a semi-passive means of providing information: the user is exposed to further details as they move their mouse over the plot. Hyperlinks are an active means of requesting further information.

This nested, hierarchical provision of information may be particularly useful for complex systematic review methodology.

It is worth noting that users should ensure they do not breach bibliographic database (and other) Terms of Use or inadvertently infringe copyright by linking to directly exported search results or including copyrighted data such as abstracts. Many providers would not count this form of transparency as outside acceptable use, but we encourage users to be certain for the resources they have used. One way to avoid this would be to provide a digitised (e.g. comma-separated text file) list of digital object identifiers for all records linked to in an interactive version of the flow diagram: this would contravene neither copyright nor Terms of Use and could be transformed into a full set of citations using freely accessible resources like CrossRef.

### Case study

We have prepared a case study that demonstrates possible interactivity that can be employed in a web-based *PRISMA2020* flow diagram (see Figure 4). The example website is available at https://driscoll.ntu.ac.uk/prisma/.

**Figure 4.**
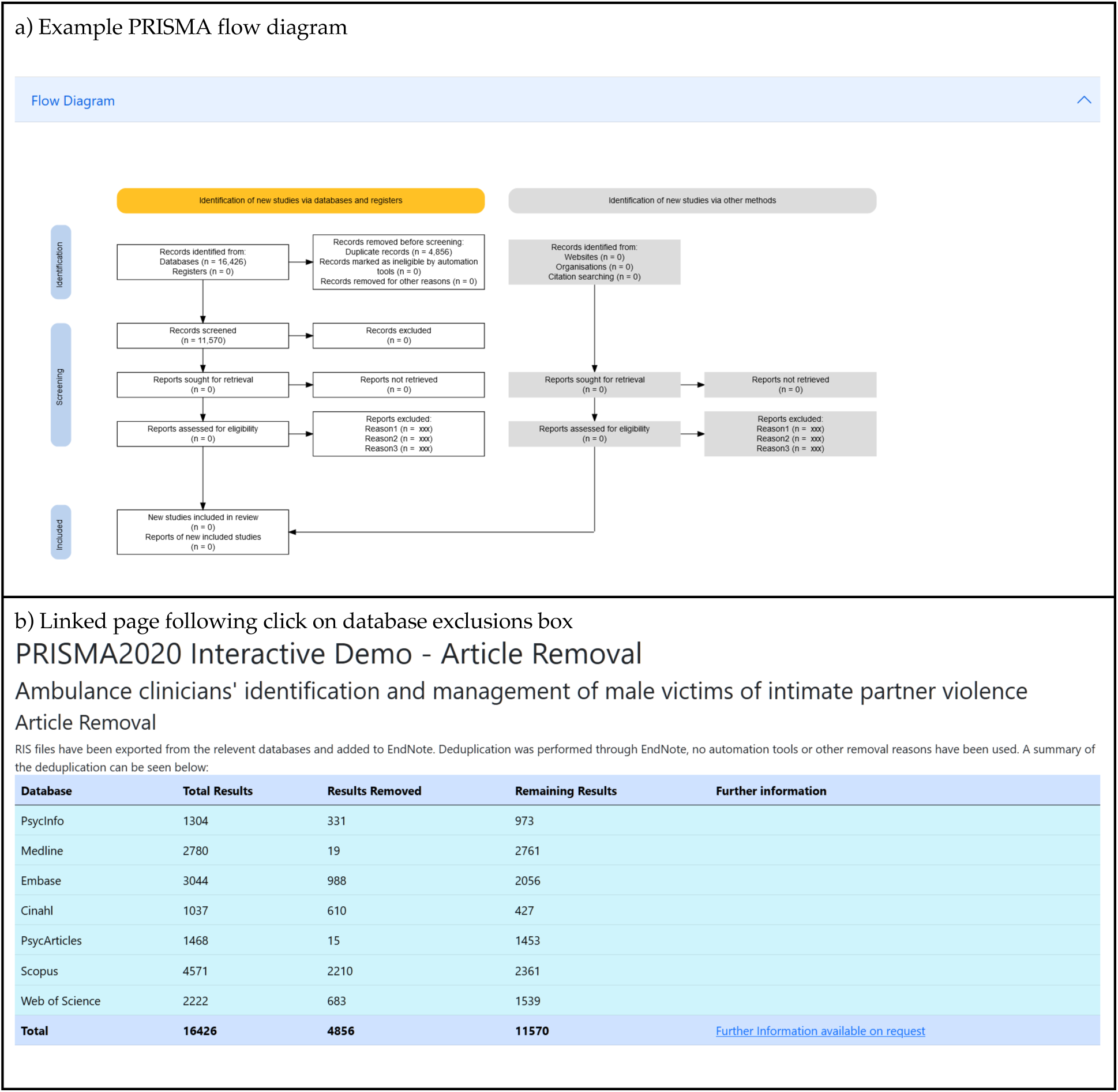
Screenshot of the case study website, showing the PRISMA flow diagram (a) and the resulting page linked to by clicking on the database exclusions box (b).

The website is based on data from an ongoing systematic review into ambulance clinician responses to adult male victims of intimate partner violence (27). The site uses a flowchart generated from this software, alongside bootstrap (https://getbootstrap.com) to make a fully interactive experience, enabling users to interrogate various aspects of the review. As the review is currently underway, the site will be updated as the review progresses.

## Discussion

The PRISMA 2020 update represents a significant development of the PRISMA statement, increasing the usability and level of detail needed in systematic reviews. The *PRISMA2020* flow diagram similarly provides a clearer and more detailed template. We have developed a user-friendly suite of tools for producing PRISMA 2020-compliant flow diagrams for users with coding experience and, importantly, for users without prior experience in coding by making use of Shiny. These free-to-use tools will make it easier to produce clear and PRISMA 2020-compliant systematic review flow diagrams. Significantly, users can also produce interactive flow diagrams for the first time, allowing readers of their reviews to smoothly and swiftly explore and navigate to further details of the methods and results of a review.

In addition, the ability to produce flow diagrams using code in a data-driven approach carries with it a number of benefits, including: facilitating Open Science (specifically Open Code); reducing the risk of transcription errors; and, opening up possibilities for reproducible documents such as executable research articles (28) and communicating the results of living systematic reviews (17).

We believe these tools will increase use of PRISMA flow diagrams, improve the compliance and quality of flow diagrams, and facilitate strong science communication of the methods and results of systematic reviews by making use of interactivity. We encourage the systematic review community to make use of these tools, and provide feedback to streamline and improve their usability and efficiency.

## Data Availability

All data described in the manuscript are available from the example website: https://driscoll.ntu.ac.uk/prisma/

## Acknowledgements

We thank Jack Wasey for his work on an R package for PRISMA 2009-compliant flow diagrams. We also thank the many users of the beta version of the R package and Shiny App for feedback instrumental in improving the user experience. This work is a project of the Evidence Synthesis Hackathon (https://www.eshackathon.org/software/PRISMA2020.html).

## Author contributions

This manuscript, the R package and the Shiny App were conceived and drafted by Neal Haddaway. Substantial code improvements were made by Luke McGuiness and Chris Pritchard. Matthew Page provided conceptual feedback. All authors contributed to the editing and revision of the manuscript, and have read and agreed to publication in its final form.

## References

1. Higgins JP, Thomas J, Chandler J, Cumpston M, Li T, Page MJ, et al. Cochrane handbook for systematic reviews of interventions: John Wiley & Sons; 2019.

2. Collaboration for Environmental Evidence. Guidelines and Standards for Evidence Synthesis in Environmental Management: Version 5.0. Project Report. 2018.

3. Moher D, Liberati A, Tetzlaff J, Altman DG, Group P. Preferred reporting items for systematic reviews and meta-analyses: the PRISMA statement. PLoS medicine. 2009;6(7):e1000097.

4. Haddaway NR, Macura B, Whaley P, Pullin AS. ROSES RepOrting standards for Systematic Evidence Syntheses: pro forma, flow-diagram and descriptive summary of the plan and conduct of environmental systematic reviews and systematic maps. Environmental Evidence. 2018;7(1):1–8.

5. Liberati A, Altman DG, Tetzlaff J, Mulrow C, Gøtzsche P, Ioannidis J. The PRISMA statement for reporting systematic and meta analyses of studies that evaluate interventions: explanation and elaboration. PLoS Medicine. 2009;6(7):1–28.

6. Moher D, Tetzlaff J, Tricco AC, Sampson M, Altman DG. Epidemiology and reporting characteristics of systematic reviews. PLoS medicine. 2007;4(3):e78.

7. Page MJ, Shamseer L, Altman DG, Tetzlaff J, Sampson M, Tricco AC, et al. Epidemiology and reporting characteristics of systematic reviews of biomedical research: a cross-sectional study. PLoS medicine. 2016;13(5):e1002028.

8. Page MJ, McKenzie JE, Bossuyt PM, Boutron I, Hoffmann TC, Mulrow CD, et al. The PRISMA 2020 statement: an updated guideline for reporting systematic reviews. Bmj. 2021;372.

9. Stovold E, Beecher D, Foxlee R, Noel-Storr A. Study flow diagrams in Cochrane systematic review updates: an adapted PRISMA flow diagram. Systematic reviews. 2014;3(1):1–5.

10. Vu-Ngoc H, Elawady SS, Mehyar GM, Abdelhamid AH, Mattar OM, Halhouli O, et al. Quality of flow diagram in systematic review and/or meta-analysis. PloS one. 2018;13(6):e0195955.

11. Kraker P, Leony D, Reinhardt W, Beham G. The case for an open science in technology enhanced learning. International Journal of Technology Enhanced Learning. 2011;3(6):643–54.

12. Pontika N, Knoth P. Open science taxonomy. 2015.

13. Wilkinson MD, Dumontier M, Aalbersberg IJ, Appleton G, Axton M, Baak A, et al. The FAIR Guiding Principles for scientific data management and stewardship. Scientific data. 2016;3(1):1–9.

14. Ehrig-Page K. Using Creative Commons Licenses and Creative Commons Licensed Works. 2020.

15. Haddaway NR. Open Synthesis: on the need for evidence synthesis to embrace Open Science. Environmental evidence. 2018;7(1):1–5.

16. Blank G, Reisdorf BC. The participatory web: A user perspective on Web 2.0. Information, Communication & Society. 2012;15(4):537–54.

17. Elliott JH, Synnot A, Turner T, Simmonds M, Akl EA, McDonald S, et al. Living systematic review: 1. Introduction—the why, what, when, and how. Journal of clinical epidemiology. 2017;91:23–30.

18. Finkler W, Leon B. The power of storytelling and video: a visual rhetoric for science communication. Journal of science communication. 2019;18(5):pA02.

19. Team RDC. R (version 3.5. 3). R: A language and environment. 2018.

20. Haddaway N, McGuinness L, Pritchard C. PRISMA2020: R package and ShinyApp for producing PRISMA 2020 compliant flow diagrams (Version 0.0.1). Zenodo. 2021.

21. Iannone R, Iannone MR. Package ‘DiagrammeR’. 2020.

22. Vaidyanathan R, Xie Y, Allaire J, Cheng J, Russell K. htmlwidgets: HTML Widgets for R. R package version 06. 2018.

23. Wickham H, Hester J, Ooms J. xml2: Parse XML. R package version 1.2. 0. 2018.

24. Dyck M, Robie J, Snelson J, Chamberlin D. XML path language (XPath) 3.1. W3C Working Draft. 2014.

25. Ooms J. rsvg: Render SVG Images into PDF, PNG, PostScript, or Bitmap Arrays R package version 1.1. R Foundation for Statistical Computing; 2017.

26. Beeley C. Web application development with R using Shiny: Packt Publishing Ltd; 2013.

27. Mackay J, Pritchard C, Jieru Yan R, Kuhn I, Barker E. Ambulance clinicians’ identification and management of male victims of intimate partner violence: a systematic review protocol PROSPERO. 2021;CRD42021253584.

28. Tsang E, Maciocci G. Welcome to a new ERA of reproducible publishing. eLIFE Labs. 2020.

